# Pharmacokinetics of Lidocaine Infusion: Optimal Dosing and Duration in ERAS Protocol

**DOI:** 10.1101/2025.08.08.25333328

**Authors:** Allen Xu, Albert Ren, Chihjen Lee

## Abstract

The Enhanced Recovery After Surgery (ERAS) protocol promotes multimodal analgesia to optimize postoperative pain control and reduce opioid consumption. To mitigate the risks of opioid-induced respiratory depression and postoperative ileus, ERAS incorporates alternative strategies, including continuous lidocaine infusion. However, excessive administration of lidocaine can cause local anesthetic systemic toxicity (LAST), potentially leading to adverse central nervous system and cardiovascular effects such as respiratory depression, seizures, coma, arrhythmia, and cardiac arrest.

This study aims to develop a pharmacokinetic model of lidocaine infusion using a one-compartment approach based on differential equations. Assuming a half-life of 2 hours and a maximum safe plasma concentration of 5 mg/kg, the model predicts lidocaine concentrations over time, estimates the optimal discontinuation point, and calculates the additional allowable bupivacaine dose for a transversus abdominis plane (TAP) block at the end of surgery to minimize the risk of LAST.

## Introduction

Postoperative pain management is an essential component of surgical care, directly impacting patient outcomes, recovery time, and overall satisfaction. The Enhanced Recovery After Surgery (ERAS) protocol has emerged as a comprehensive, evidence-based approach that emphasizes multimodal analgesia to reduce opioid use and its associated complications [Ljungqvist et al., 2017]. Among these complications, opioid-induced respiratory depression and postoperative ileus represent serious risks, necessitating the development of alternative analgesic strategies [Bateman et al., 2021].

Intravenous lidocaine has garnered increasing attention within ERAS protocols for its analgesic, anti-inflammatory, and opioid-sparing properties. When administered via continuous infusion, lidocaine can provide effective pain relief without the sedative and respiratory side effects associated with opioids [Beaussier et al., 2018]. However, its narrow therapeutic window requires careful monitoring, as excessive plasma concentrations may result in local anesthetic systemic toxicity (LAST). Clinical manifestations of LAST range from mild neurological symptoms to severe central nervous system and cardiovascular depression, including seizures, loss of consciousness, coma, arrhythmia, and cardiac arrest [Dickerson et al., 2014].

Despite its clinical advantages, the safe administration of lidocaine infusion remains challenging due to inter-patient variability in metabolism and elimination. To address this, a pharmacokinetic based strategy is needed to guide clinical dosing and reduce the risk of toxicity. This study employs a one-compartment pharmacokinetic model and uses differential equations to simulate lidocaine concentration over time. Assuming a two-hour half-life and a maximum safe dose of 5 mg/kg, the model estimates the optimal time to discontinue infusion and predicts the residual lidocaine levels. Furthermore, it enables clinicians to calculate the maximum additional safe dose of bupivacaine for a transversus abdominis plane (TAP) block at the conclusion of surgery, thereby improving the safe co-administration of lidocaine and bupivacaine.

## Methods

### Deriving the rate constant

First, the rate constant for lidocaine must be determined. The following equation is used, where 2 represents the half-life of lidocaine (in hours) and r denotes the rate constant.

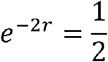

To solve the equation above, we take the natural logarithm on both sides.

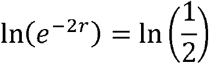

Here *ln* and *e* cancel each other out.

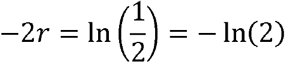

Therefore.

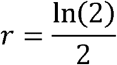

### Setting up the differential equation

The change in lidocaine amount over time can be described in the following differential equation, where 2 is the infusion rate in mg per hour, 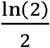 is the rate constant derived from half-life (as shown earlier), *y* is the lidocaine amount in the system, and *t* is time in hours.

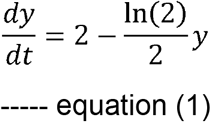

As indicated by the second part of the equation, the rate of decline is directly proportional to the total lidocaine amount in the system.

## Results

### The maximal dose achieved at the steady state

At the steady state, the rate of change of the level of lidocaine would be 0. Therefore, to solve the steady state lidocaine drug level, we can set 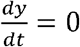 and solve for y:

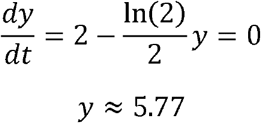

This result indicates that, if lidocaine is continuously infused at a constant rate of 2mg/kg/hr, the amount will asymptotically approach a steady-state level of approximately 5.77 mg/kg over time.

### Solving the differential equation

After solving Equation (1), we get the following equation, which describes the amount of lidocaine in the system as a function of time.

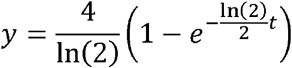

The general solution is shown below.

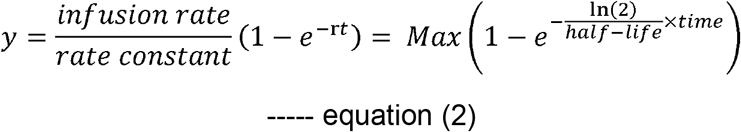

### When lidocaine level reaches 4 mg/kg

To ensure safety, the infusion should be stopped before lidocaine reaches its maximum allowable concentration. To determine when the lidocaine level will reach 4 mg/kg, we set y = 4, as shown below.

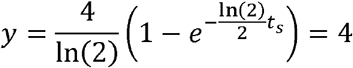

Solving for *t*_*s*_(the stop time), we find *t*_*s*_is approximately 3.41 h

Therefore, when we stop the lidocaine infusion at **3.41 hours**(approximately 3 hours and 25 minutes), the level of lidocaine in the system is roughly **4 mg/kg**.

### When lidocaine level reaches 5 mg/kg

By setting *y*= 5, we can calculate the time at which the lidocaine level reaches 5 mg/kg using the same equation:

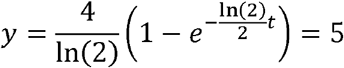

Solving for time *t*, we obtain an estimated value of approximately 5.81 hours.

This means that if lidocaine infusion continues at a constant rate of 2 mg/kg/hr, the drug level in the system will reach about **5 mg/kg**at around **5.81 hours**(5 hours and 49 minutes).

### Setting up the exponential decay of lidocaine

Suppose lidocaine infusion is stopped at 3.41 hours (3 hours and 25 minutes). Now the amount of lidocaine will decline exponentially according to the following equation:

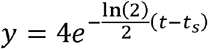

In the equation above, the value 4 represents the initial amount of 4 mg/kg at the time the infusion is stopped, 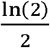 is the rate constant, and (t ‐ t_⍰_) denotes the time elapsed since the infusion was discontinued.

### Graphing the lidocaine dosage over time

With an infusion rate of 2 mg/kg/hr, the lidocaine level is modeled using a one-compartment pharmacokinetic approach. The following graph illustrates the “hyperbolic” rise in lidocaine levels, the time required to reach the threshold of 4 mg/kg, and the subsequent decline over four hours (equivalent to two half-lives). This represents a total time course of 7 hours and 25 minutes prior to the TAP block. Please note that the surgery may finish earlier, and the amount of lidocaine remaining should be estimated accordingly.

**Fig. 1.**
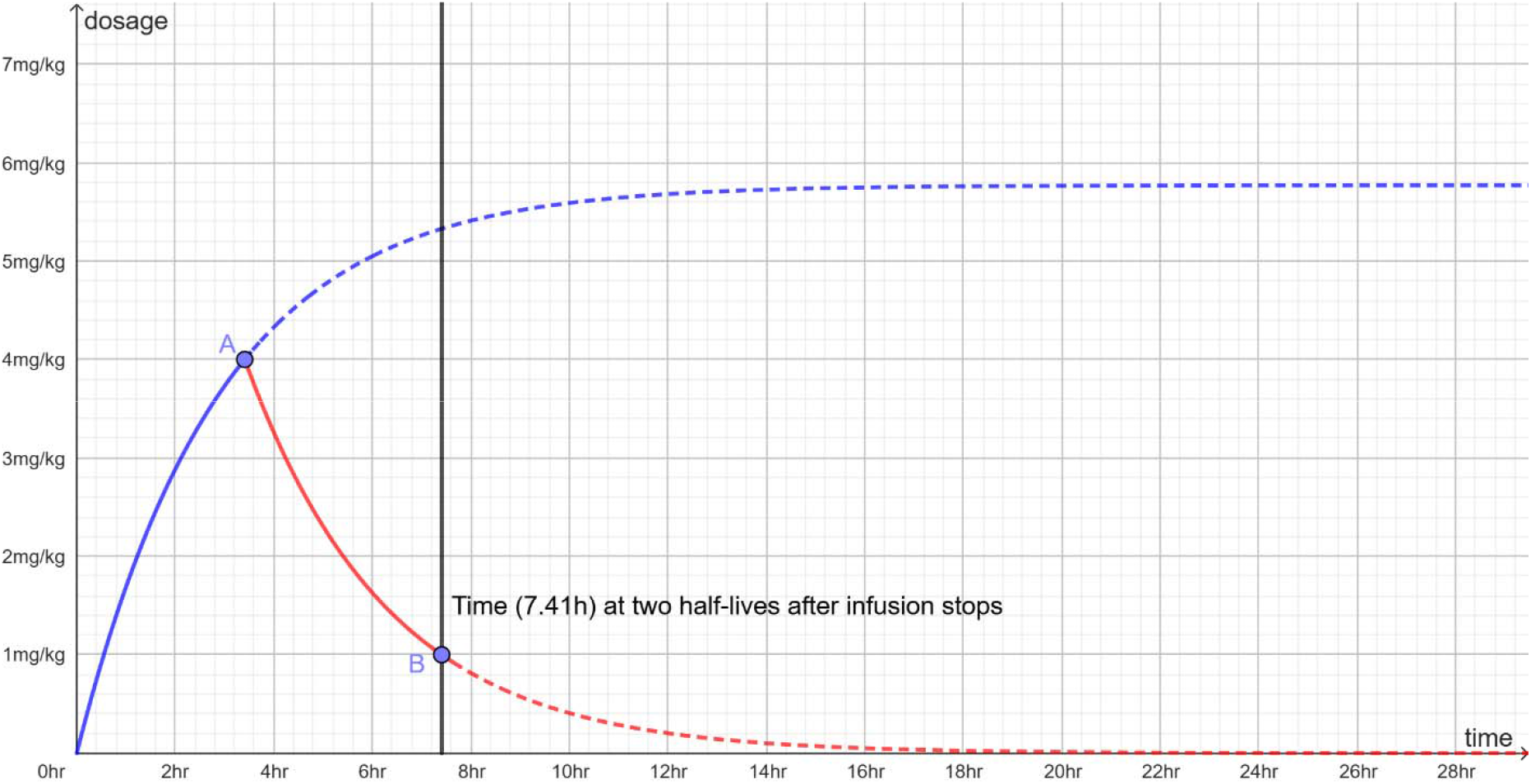
The lidocaine dose increases “hyperbolically”, reaching 4 mg/kg at 3.41 hours (point A). At that point the infusion is discontinued. Given the drug’s half-life, the dose is expected to fall to 1 mg/kg four hours (two half-lives) later, representing a total time of 7.41 hours (point B).

## Discussion

### ERAS protocol for bowel surgery

As a postoperative recovery strategy, the Enhanced Recovery After Surgery (ERAS) protocol incorporates a range of evidence-based interventions aimed at accelerating recovery and improving patient outcomes. Among these interventions, intravenous lidocaine infusion serves as an effective component of multimodal analgesia [Ljungqvist et al., 2017]. Accurate pharmacokinetic modeling of lidocaine is essential to ensure its safe and effective integration with other ERAS practices, particularly those involving additional anesthetic agents such as bupivacaine. Understanding the pharmacodynamic interactions and clearance profiles of these agents enables optimized dosing strategies and minimizes the risk of systemic toxicity.

### Lidocaine pharmacokinetics

Lidocaine is a commonly used amide-type local anesthetic with well-characterized pharmacokinetics. Its maximum allowable level typically lies between 4.5 and 5 mg/kg, depending on the route of administration. When given intravenously (IV)—as is common in continuous infusions for analgesia—the systemic absorption is immediate, and plasma concentrations must be carefully monitored due to the narrow therapeutic window. In contrast, subcutaneous or local infiltration results in slower absorption and lower peak plasma concentrations, allowing for a higher maximum safe dose (up to 7 mg/kg when combined with epinephrine) [Masic et al., 2018]. Without epinephrine, the time to peak plasma concentration is 20–45 minutes, whereas with epinephrine, it extends to 60–90 minutes. Using equation (2), with a dose of 5 mg/kg, a half-life of 2 hours, a peak time at 30 minutes and the assumption of total absorption, we find the peak concentration to be 4.6 mg/kg—only slightly lower than the total given dose.

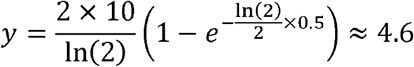

Of note, to prioritize patient safety and minimize the risk of systemic toxicity, this study conservatively sets the target systemic dose at 4 mg/kg.

This pharmacokinetic property is particularly relevant when determining safe intervals between lidocaine administration and subsequent anesthetic procedures, such as transversus abdominis plane (TAP) blocks involving bupivacaine. Understanding the systemic exposure from prior lidocaine dosing—especially via IV—ensures that cumulative local anesthetic levels remain below toxicity thresholds, promoting safer perioperative care.

### Ideal body weight and real body weight

In the context of continuous lidocaine infusion as part of Enhanced Recovery After Surgery (ERAS) protocols, precise dosing is paramount to maximizing analgesic benefits while minimizing systemic toxicity. Given lidocaine’s pharmacokinetic profile— characterized by limited distribution into adipose tissue—dosing based on ideal body weight (IBW) is recommended over actual body weight (ABW) for infusion calculations [Cheymol et al., 1993; Siddiqui et al., 2025]. This approach better aligns with the drug’s effective volume of distribution, reducing the risk of overdosing, particularly in obese patients undergoing surgery.

The commonly used formulas for calculating IBW are as follows [Peterson et al., 2016]:

- For men:

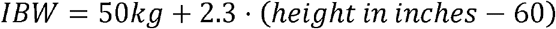
- For women:

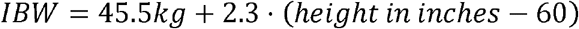

For patients with obesity, where actual body weight (ABW) exceeds IBW by more than 30%, an adjusted body weight (AdjBW) may be considered to better approximate drug distribution [Peterson et al., 2016]:

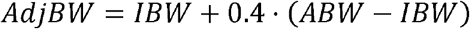

While IBW remains the preferred standard for initial dosing and infusion rate calculations, using AdjBW for prolonged infusions can be appropriate, provided that close clinical monitoring and, if available, therapeutic drug level assessments are performed to mitigate toxicity risks.

Applying these weight-based dosing strategies within ERAS protocols promotes individualized and safer lidocaine infusion management, optimizing analgesic efficacy while minimizing systemic toxicity. Further refinement of pharmacokinetic models tailored to varied body compositions will enhance dosing accuracy and improve clinical outcomes in continuous lidocaine therapy.

### Lidocaine - bupivacaine dose conversion

**Table 1.** This table shows the difference between the maximum dosage of each local anesthetic with and without epinephrine [Chietero et al.].

| Drug | Onset | Max dose (mg/kg) | Max dose with Epi (mg/kg) |
| --- | --- | --- | --- |
| Lidocaine | Rapid | 4.5 | 7 |
| Mepivacaine | Medium | 5 | 7 |
| Bupivacaine | Slow | 2.5 | 3 |
| Ropivacaine | Slow | 4 | N/A |
| Tetracaine | Slow | 1.5 | N/A |
| Chlorprocaine | Rapid | 10 | 15 |

Unlike lidocaine, bupivacaine is more potent and has a longer duration of action. Consequently, a lower dose of bupivacaine is sufficient to reach toxic levels [Smrkolj et al., 2023]. The maximum safe dose for bupivacaine is approximately 2.5 mg/kg, compared to 4.5 mg/kg for lidocaine. Therefore, estimating the appropriate bupivacaine dose following lidocaine infusion requires careful adjustment based on residual lidocaine levels to avoid exceeding the combined toxicity threshold. As our example shows, at the end of a 7.5-hour surgery, approximately 1 mg/kg of lidocaine remains in the system. This suggests that a maximum additional bupivacaine dose of 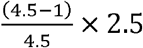 mg/kg ≈ 1.94 mg/kg can be safely administered, assuming the maximum dose of lidocaine is 4.5 mg/kg.

### Recommendation on lidocaine infusion

The results of this study suggest that the recommended stopping time for lidocaine infusion is 3.41 hours, at which point the drug level reaches 4 mg/kg. Approximately 4 hours after discontinuation, the lidocaine level declines to around 1 mg/kg. This timeline provides a basis for calculating the appropriate dosage for subsequent bupivacaine administration in a TAP block. However, since the duration of surgery may vary, the remaining lidocaine level and the maximum allowable bupivacaine dose should be recalculated accordingly.

### The combined use with epinephrine

The local vasoconstriction caused by epinephrine reduces blood flow at the injection site, allowing lidocaine and bupivacaine to be absorbed more slowly and increasing the duration of anesthesia significantly. As a result, combining epinephrine with these local anesthetics raises the maximum safe dosage [Garutti et al., 2008]. Due to differences in potency and toxicity profiles, the extent of this increase is different between the two drugs. For lidocaine, the maximum recommended dose increases from 5 mg/kg to 7 mg/kg when combined with epinephrine. In contrast, bupivacaine’s maximum dose increases more modestly, from 2.5 mg/kg to 3 mg/kg.

### Patients with different pharmacokinetic profiles

The half-life of local anesthetics can vary significantly depending on individual patient circumstances. Factors such as concurrent medication use and overall health status can influence the drug’s metabolism and elimination. For example, patients with **liver failure**may exhibit a prolonged half-life of approximately **343 minutes**, while those with congestive heart failure (CHF) or renal impairment may have slightly prolonged half-lives of around 136 minutes and 133 minutes, respectively.

To ensure patient safety, it is essential to consider lidocaine’s half-life, which can be expressed by the following formula:

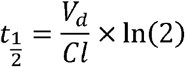

where *v*_*d*_ represents the volume of distribution (L), and *cl* represents clearance (L/hr). Please note that, theoretically, a patient with a higher volume of distribution (*v*_*d*_) or slower clearance (*cl*) would have a longer half-life. However, the effects of *v*_*d*_ and *cl* may counterbalance each other, making accurate estimation of half-life more challenging. The actual half-life is best determined by measuring drug concentrations in the patient’s body over time.

### Lidocaine concentration and toxicity

The risk of lidocaine-induced systemic toxicity is closely related to its plasma concentration. While the generally accepted maximum dose of lidocaine without epinephrine is 5 mg/kg [Tucker et al., 1979], this study adopts a more conservative threshold of 4 mg/kg to enhance patient safety during continuous infusion in the ERAS protocol. Clinical data indicate that central nervous system (**CNS**) **toxicity**typically begins to manifest at concentrations around **5 to 7 µg/mL**, with symptoms such as tinnitus, dizziness, perioral numbness, metallic taste, and tremors. As levels rise beyond 7 µg/mL, the likelihood of severe adverse effects, including seizures and loss of consciousness, increases significantly. The **Lethal**level is generally associated with plasma concentrations exceeding **10 to 12 µg/mL**, at which point profound CNS depression, respiratory arrest, hypotension, and cardiovascular collapse may occur [Tucker et al., 1979]. These thresholds underscore the importance of carefully managing infusion rates and patient-specific dosing parameters—such as ideal body weight and hepatic function—to prevent accumulation and systemic toxicity.

To ensure safe and effective anesthetic management, it is essential to distinguish between the drug’s plasma concentration and its total amount in the body. While concentration (mg/L) reflects the measurable level of lidocaine in the bloodstream, the total amount (mg) represents the absolute quantity present in the patient’s body. These two parameters are connected through the volume of distribution (*v*_*d*_), expressed in liters per kilogram (L/kg), and calculated using the equation:

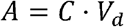

where:

- *A*is the total amount of lidocaine in the body (mg),
- *c*is the plasma concentration (mg/L), and
- *v*_*d*_is the volume of distribution (L/kg).

Roughly, the volume of distribution for lidocaine ranges from 0.7 to 1.5 L/kg. Within this range, the total amount of lidocaine in the body can be converted to the corresponding plasma concentration. [Foong et al., 2024]:

#### Toxic concentration

Suppose toxic lidocaine concentration is 6 μ*g*per ml.

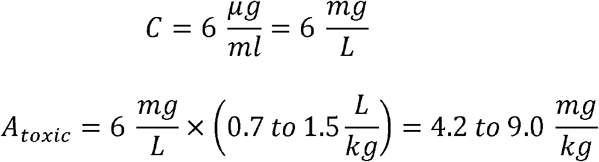

This calculation shows that the toxic level can be reached readily with an infusion rate of 2 mg/kg/hour.

#### Lethal concentration

Suppose the lethal concentration is 10 μ*g*per ml.

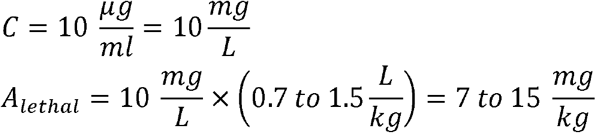

This level can be reached, for instance, in liver patients who have a half-life of 343 minutes (5.7 hours) as shown below.

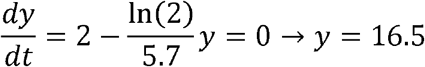

In liver patients, non-stop lidocaine infusion may be dangerous due to an increased volume of distribution and reduced clearance.

We can find out the time to stop by solving y and set *y*= 4.

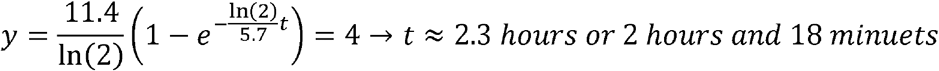

This calculation indicates that, in patients with **liver failure**, the infusion should be discontinued after **2 hours and 18 minutes**.

## Data Availability

All data produced in the present work are contained in the manuscript

## Author Contributions

Allen Xu: writing original draft, calculation, visualization, review and editing, proofreading.

Albert Ren: review and editing, proofreading.

Chihjen Lee: conceptualization, methodology, supervision, visualization, review and editing, proofreading.

